# The relationship between serotonin transporter occupancy and extracellular serotonin concentration is hyperbolic, not linear: implications for safely tapering antidepressants

**DOI:** 10.64898/2026.06.09.26355019

**Authors:** Daniel Cohrs, Bryan Shapiro

**Affiliations:** Department of Psychiatry and Behavioral Sciences, Keck School of Medicine, University of Southern California, Los Angeles, CA, USA; Department of Psychiatry and Human Behavior, UC Irvine Medical Center, Orange, CA, USA

**Keywords:** Serotonin, Serotonin reuptake inhibitors, Antidepressants, Antidepressant withdrawal, Hyperbolic tapering, Deprescribing

## Abstract

**Background:** Hyperbolic tapering is an increasingly recognized approach for discontinuing serotonin reuptake inhibitor (SRI) antidepressants that involves non-linear dose reductions with equal stepwise reductions in serotonin transporter (SERT) occupancy to mitigate withdrawal symptoms. Its theoretical basis is the hyperbolic relationship between SRI dose and SERT occupancy reported in radioligand imaging studies. Hyperbolic tapering implicitly assumes that changes in SERT occupancy approximate changes in biologic effect and withdrawal risk. Because SERT occupancy plateaus across the therapeutic dose range of SRIs, this framework predicts relatively small biologic effects and withdrawal risk within this range. However, SERT occupancy influences serotonergic activity only indirectly via its effects on extracellular serotonin concentrations, and the relationship between these two variables is poorly characterized.

**Methods:** We developed a two-pathway clearance model derived from mass-action kinetics to evaluate the steady-state relationship between SERT occupancy and extracellular serotonin concentrations under chronic SRI treatment.

**Results:** Our analysis indicates that serotonin concentrations increase hyperbolically as transporter occupancy increases, suggesting that biologically meaningful differences in serotonergic signaling persist across the therapeutic dose range of SRIs despite plateauing occupancy.

**Conclusions:** Our model predicts a hyperbolic relationship between SERT occupancy and extracellular serotonin concentrations, suggesting that changes in occupancy may not map proportionally onto serotonergic effect. These findings provide a potential mechanistic explanation for dose-dependent clinical effects of SRIs despite plateauing transporter occupancy and generate testable hypotheses regarding antidepressant tapering strategies. Empirical validation is warranted.

## 1. Introduction

Serotonin reuptake inhibitors (SRIs) (e.g. selective serotonin reuptake inhibitors, serotonin-norepinephrine reuptake inhibitors, tricyclic antidepressants) are the most widely prescribed pharmacologic treatments for depressive and anxiety disorders, with a median duration of use of 2-5 years in the US and UK, respectively (Kendrick, 2021; Ward et al., 2025). Withdrawal symptoms are common when discontinuing SRI treatment, and consist of a constellation of cognitive, affective and somatic disturbances (Horowitz and Taylor, 2019). While withdrawal symptoms are more commonly mild and self-limiting in short-term (<6 months) users, they are more likely to be severe, prolonged and associated with significant functional impairment in long-term users. (Taylor and Horowitz, 2024)

For the many patients who experience withdrawal symptoms using linear tapering (i.e. tapering to the minimum available dose using standard dose formulations before discontinuation), hyperbolic tapering has gained prominence as a preferred approach for SRI discontinuation and has been incorporated into formal clinical guidance in the UK and Australia. (Horowitz and Taylor, 2019; Horowitz and Wilcock, 2025) This approach is grounded in the hyperbolic relationship between SRI dose and serotonin transporter (SERT) occupancy demonstrated in radioligand imaging studies (Figure 1). (Meyer et al., 2004)

**Figure 1.**
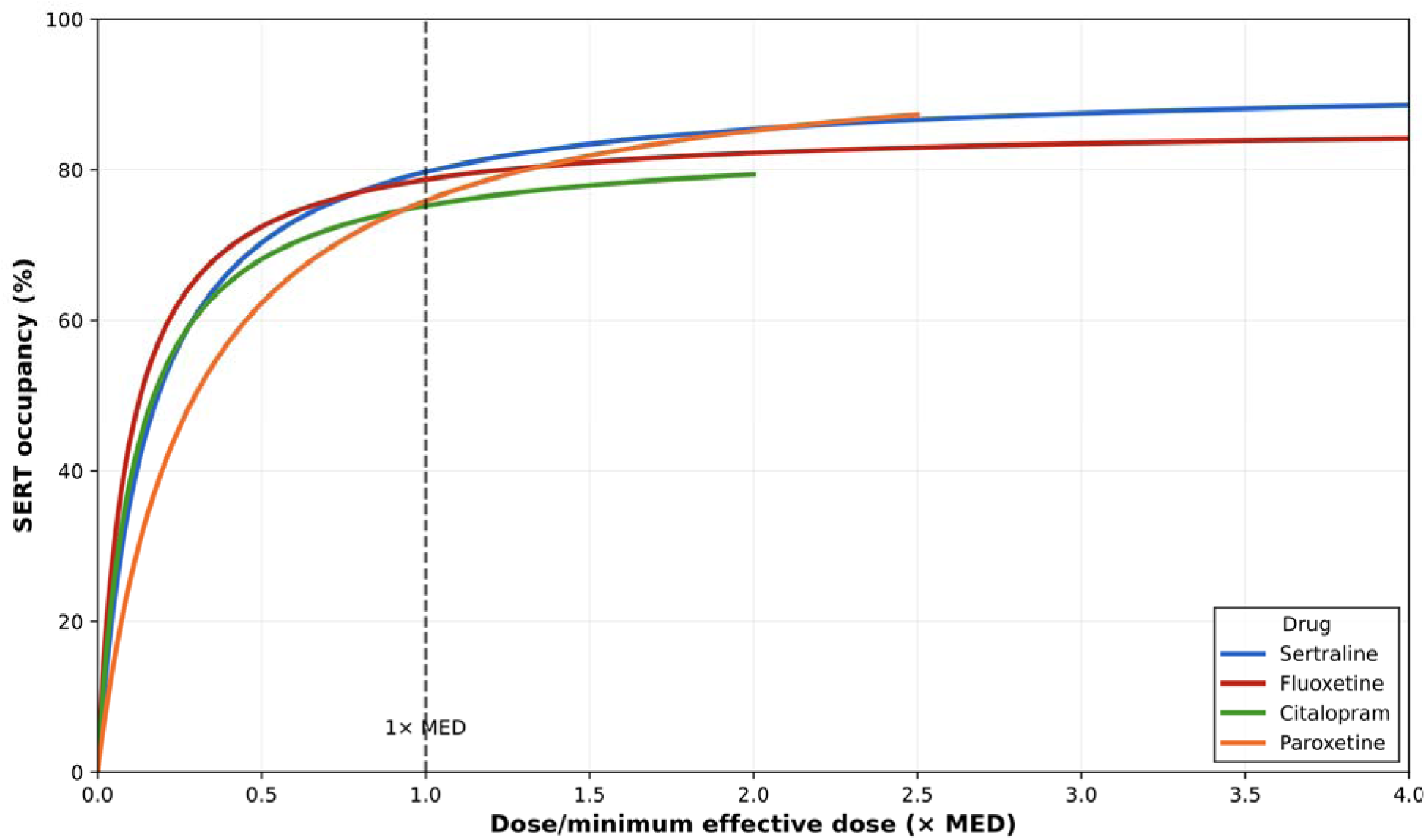
Dose-occupancy curves for SSRI antidepressants, normalized to the U.S. Food and Drug Administration (FDA) minimum effective dose for the treatment of major depressive disorder.(Sørensen et al., 2022) Curves are truncated at the F.D.A. maximum recommended doses. Abbreviations: SSRI=selective serotonin reuptake inhibitor

Because the dose-occupancy curve for all SRIs plateaus in the therapeutic dose range (Figure 1), hyperbolic tapering involves larger dose reductions above the minimum therapeutic dose, and relatively smaller dose reductions below this threshold. (Shapiro, 2018) However, many patients experience withdrawal symptoms when tapering within the therapeutic dose range, despite minimal changes in SERT occupancy—observations that appear difficult to reconcile within an occupancy-based framework alone.

Dose-dependent clinical effects of SRIs are also difficult to fully reconcile within an occupancy-based model. A recent meta-analysis found significantly higher dropout rates due to adverse effects with high-dose selective serotonin reuptake inhibitor (SSRI) treatment compared to low- or medium-dose treatment with effects distributed proportionally across dose groups,(Braun et al., 2020) confirming findings from prior meta-analyses.(Furukawa et al., 2019; Jakubovski et al., 2016) Clinical trial data submitted to the FDA for escitalopram—the only SSRI for which adverse reactions were reported separately by dose rather than pooled—demonstrate linear dose-dependent increases in rates of adverse effects.(US Food and Drug Administration, 2024) While there is mixed evidence of a dose-response relationship for SSRIs in treating major depressive disorder,(Furukawa et al., 2019; Jakubovski et al., 2016) there is such evidence for the treatment of anxiety disorders(Jakubovski et al., 2019) and for obsessive-compulsive disorder (OCD). (Bloch et al., 2010)

Relative to SERT occupancy, the more proximal driver of serotonergic signaling is extracellular serotonin concentrations. Hyperbolic tapering frameworks implicitly assume that changes in SERT occupancy approximate changes in serotonergic effect. However, observations of withdrawal symptoms and dose-dependent clinical effects despite plateauing occupancy raise the possibility that the relationship between SERT occupancy and extracellular serotonin concentrations may be nonlinear. To explore this possibility, we developed a two-pathway clearance model linking SERT occupancy to extracellular serotonin concentrations under chronic SRI treatment.

## 2. Methods

Best *et al.(Best et al., 2010)* developed a mathematical model of serotonin (5-HT) synthesis, release, and reuptake in which C_5-HT_ is determined by the balance between vesicular release into the extracellular space and multiple clearance processes. In that framework, C_5-HT_ dynamics are governed by a general mass-balance equation (Figure 2).

**Figure 2.**
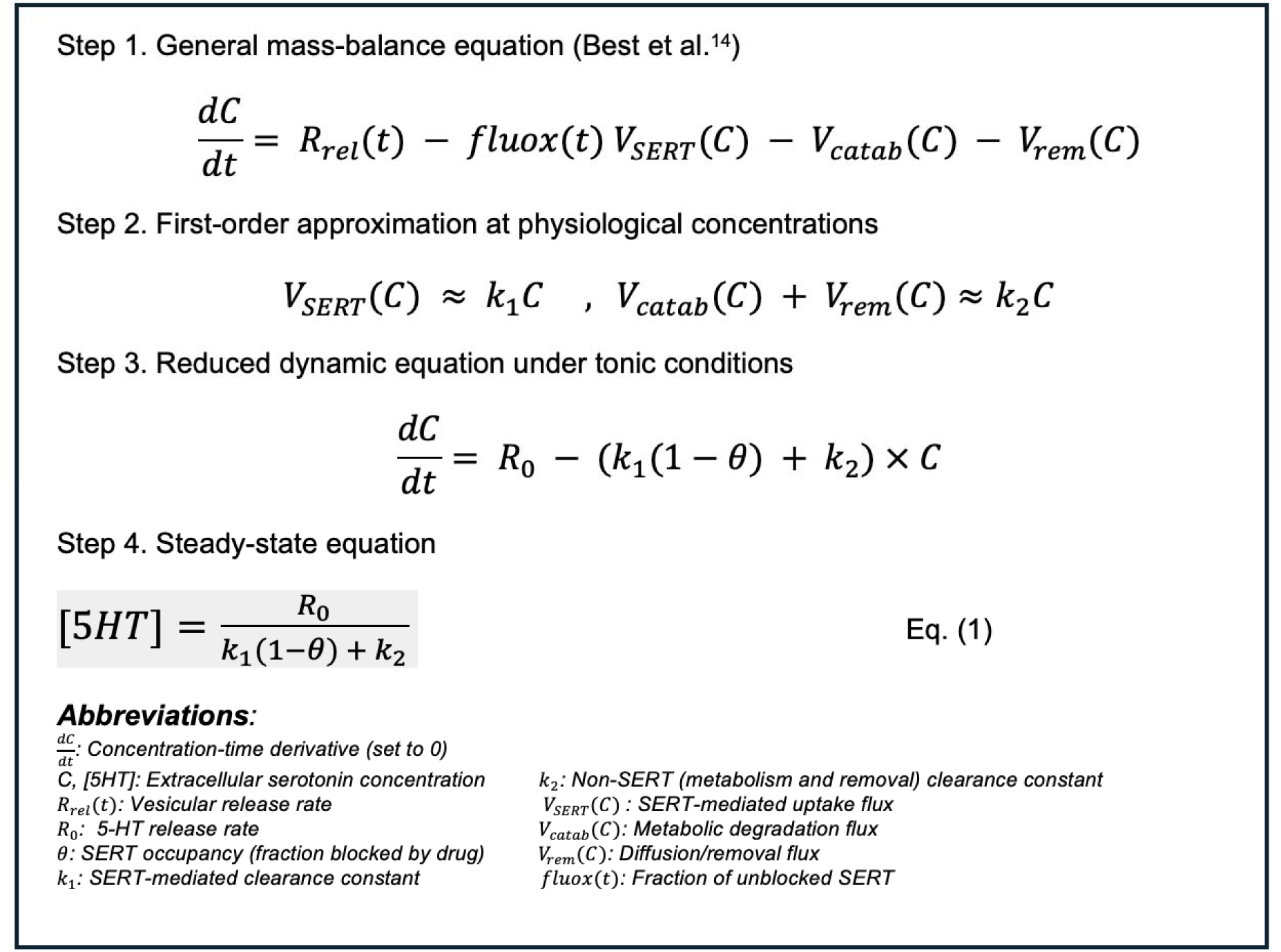
Derivation of a two-pathway mass-action clearance model for extracellular serotonin. The original mass-balance equation described by Best et al.(Best et al., 2010) is progressively simplified under physiological assumptions to yield the steady-state relationship presented in Equation (1).

At physiological concentrations, C_5-HT_ levels are well below the Michaelis constant for SERT,(Hagan et al., 2010) and cytoplasmic 5-HT concentrations are similarly well below the Michaelis constant for monoamine oxidase (MAO). (Fitzgerald et al., 1990; Verleysdonk et al., 2004) Under these conditions, both SERT-mediated reuptake and non-SERT clearance processes can be approximated as first-order functions of C_5-HT_, while passive diffusion is inherently first-order. These assumptions allow the original equation to be simplified without altering the qualitative shape of the occupancy–serotonin relationship.

At steady state, vesicular release is represented as a constant release term, R_0_, corresponding to steady 5-HT input into the extracellular space. Transporter occupancy by an SRI is represented by θ, the fraction of SERTs blocked, such that the fraction of unblocked transporters is 1−θ, consistent with the notation used by Best *et al. (Best et al., 2010)*

At steady state (dC/dt=0), this formulation yields an expression (Equation 1) that provides a parsimonious mapping from SERT occupancy to relative changes in C_5-HT_.

Although experimental values for K_1_ (SERT-mediated C_5-HT_ clearance) and K_2_ (non-SERT clearance) vary, only their relative magnitude is needed to determine the rate of growth of C_5-HT_ as a function of SERT occupancy, expressed as the K_1_:K_2_ ratio. Established ratios vary somewhat by brain region,(Tao et al., 2000) with higher K_1_:K_2_ ratios accentuating increases in C_5-HT_ as a function of SERT occupancy (Figure 3).

**Figure 3:**
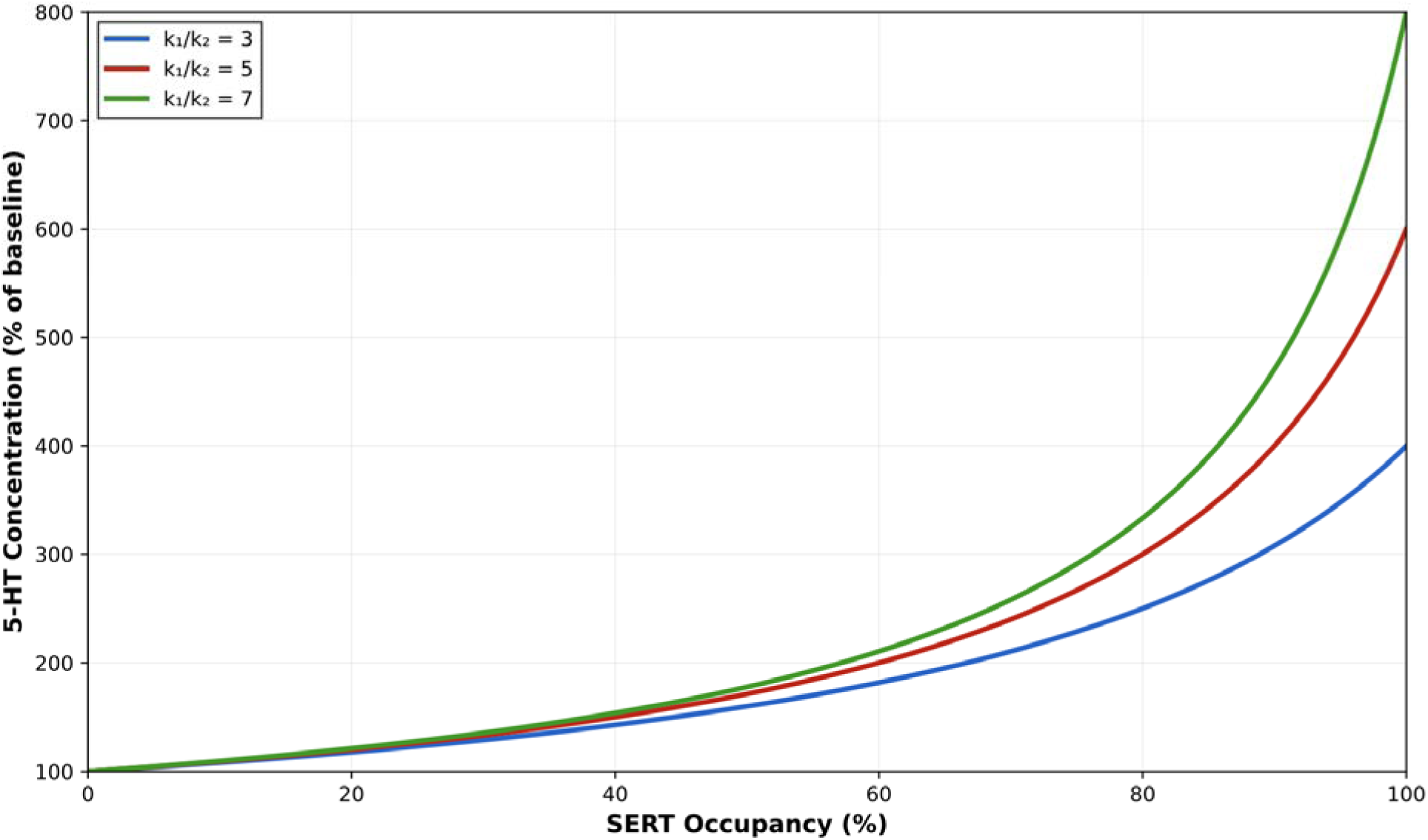
The relationship between extracellular serotonin concentration and SERT occupancy, modeled with varying K_1_:K_2_ ratios (ratio of 5:1 was used in our model) Abbreviations: SERT= serotonin transporter.

Our model was calibrated to match both whole-brain and frontal cortical estimates of C_5-HT_ elevations following chronic SRI treatment. Frontal cortical estimates were included because this region has the most robust region-specific data regarding chronic SRI-induced C_5-HT_ elevation and is thought to be particularly relevant to SRI therapeutic effects, in part because delayed serotonergic elevation in this region parallels the delayed onset of therapeutic effects. (Fritze et al., 2017) Frontal cortical microdialysis studies demonstrate consistent elevations to ∼300–350% of baseline following chronic SRI treatment (Fritze et al., 2017), while whole-brain cerebrospinal fluid estimates in primates show similar elevations. (Anderson et al., 2005) We therefore calibrated our model to a 5:1 K_1_:K_2_ ratio to reproduce elevations to 300-350% of baseline at therapeutic doses. Other brain regions were not modeled separately because available data remain comparatively sparse, although existing evidence suggests substantial regional variability in the magnitude of C_5-HT_ elevation following chronic SRI treatment (Fritze et al., 2017)

## 3. Results

Table 1 models the relationship between SERT occupancy (conventionally abbreviated “RO” to refer to receptor occupancy more broadly) and C_5-HT_ based on our two-pathway kinetic equation (red line represents the 5:1 K_1_:K_2_ ratio chosen for our model). The model predicts a hyperbolic relationship, reaching a maximal 6x of baseline at 100% RO, consistent with SERT knockout studies reporting elevations to approximately 5–9x of baseline in C_5-HT_ in serotonergic projection regions (e.g., frontal cortex, hippocampus, substantia nigra), depending on brain region and experimental conditions.(Fabre et al., 2000; Gobbi et al., 2001; Homberg et al., 2007) Substantial increases in C_5-HT_ are observed at ≥ 70-80% SERT occupancy, which corresponds to the typical therapeutic dosage range of SRI antidepressants. Smaller, but non-negligible stepwise changes in C_5-HT_ as a function of dose are observed at low ROs.

**Table 1:** Extracellular serotonin concentration as a function of SERT occupancy (RO), calculated using Equation 1. K_1_:K_2_ ratio has been set to 5:1, and C_0_ is set to 1 for illustrative purposes. Abbreviations: 5-HT=serotonin

| RO (0–50%) | 5-HT (% Base) | Δ 5-HT (%) | RO (55–100%) | 5-HT (% Base) | Δ 5-HT (%) |
| --- | --- | --- | --- | --- | --- |
| 0% | 100.0 | — | 55% | 184.6 | 7.70 |
| 5% | 104.3 | 4.30 | 60% | 200.0 | 8.34 |
| 10% | 109.1 | 4.60 | 65% | 218.2 | 9.10 |
| 15% | 114.3 | 4.77 | 70% | 240.0 | 9.99 |
| 20% | 120.0 | 4.99 | 75% | 266.7 | 11.13 |
| 25% | 126.3 | 5.25 | 80% | 300.0 | 12.49 |
| 30% | 133.3 | 5.54 | 85% | 342.9 | 14.30 |
| 35% | 141.2 | 5.93 | 90% | 400.0 | 16.65 |
| 40% | 150.0 | 6.23 | 95% | 480.0 | 20.00 |
| 45% | 160.0 | 6.67 | 100% | 600.0 | 25.00 |
| 50% | 171.4 | 7.13 |  |  |  |

The nonlinear relationship we observed applies to steady-state systems in which clearance capacity (V_max_) is linearly reduced against a sufficiently constrained input, yielding an inverse relationship between clearance capacity and concentration (C ∝ 1/V_max_). (Rowland and Tozer, 2019) In the present model (Eq. 1), V_max_ is proportional to the fraction of unblocked transporters (1 − θ). As transporter availability (1 − θ) decreases, equivalent increases in occupancy produce increasingly larger changes in C_5-HT_. This yields a hyperbolic relationship between SERT occupancy and extracellular serotonin concentrations, consistent with the general form of a hyperbolic clearance function (y=A/(B-x)), where C_5HT_=R_0_/(K_1_(1−θ) + K_2_).

## 4. Discussion

Our two-pathway kinetic model predicts a hyperbolic relationship between SERT occupancy and extracellular serotonin concentrations, suggesting that substantial changes in serotonergic signaling may persist across the therapeutic dose range despite plateauing occupancy. This finding provides a potential mechanistic explanation for clinical observations of withdrawal symptoms occurring in the therapeutic range, dose-dependent side effects attributed to increased 5-HT receptor stimulation,(Ferguson, 2001) and dose-response trends for anxiety disorders and OCD. (Jakubovski et al., 2019),(Bloch et al., 2010)

Extending our model findings to a dose-effect analysis using established dose-occupancy data for sertraline (Figure 4) reveals substantial, though diminishing, increases in C_5-HT_ across the therapeutic dosage range—an 84.3% absolute and a 28% relative increase. At very low doses (e.g. corresponding to <20% SERT occupancy), hyperbolic tapering models predict a heightened risk of withdrawal symptoms because the SRI dose-occupancy curve is very steep in this range. Our dose-effect model shows a less pronounced, although still steep, decline in C_5-HT_ at low occupancies (Figure 4), raising the possibility that taper end phases could be less gradual than current occupancy-based models predict.

**Figure 4:**
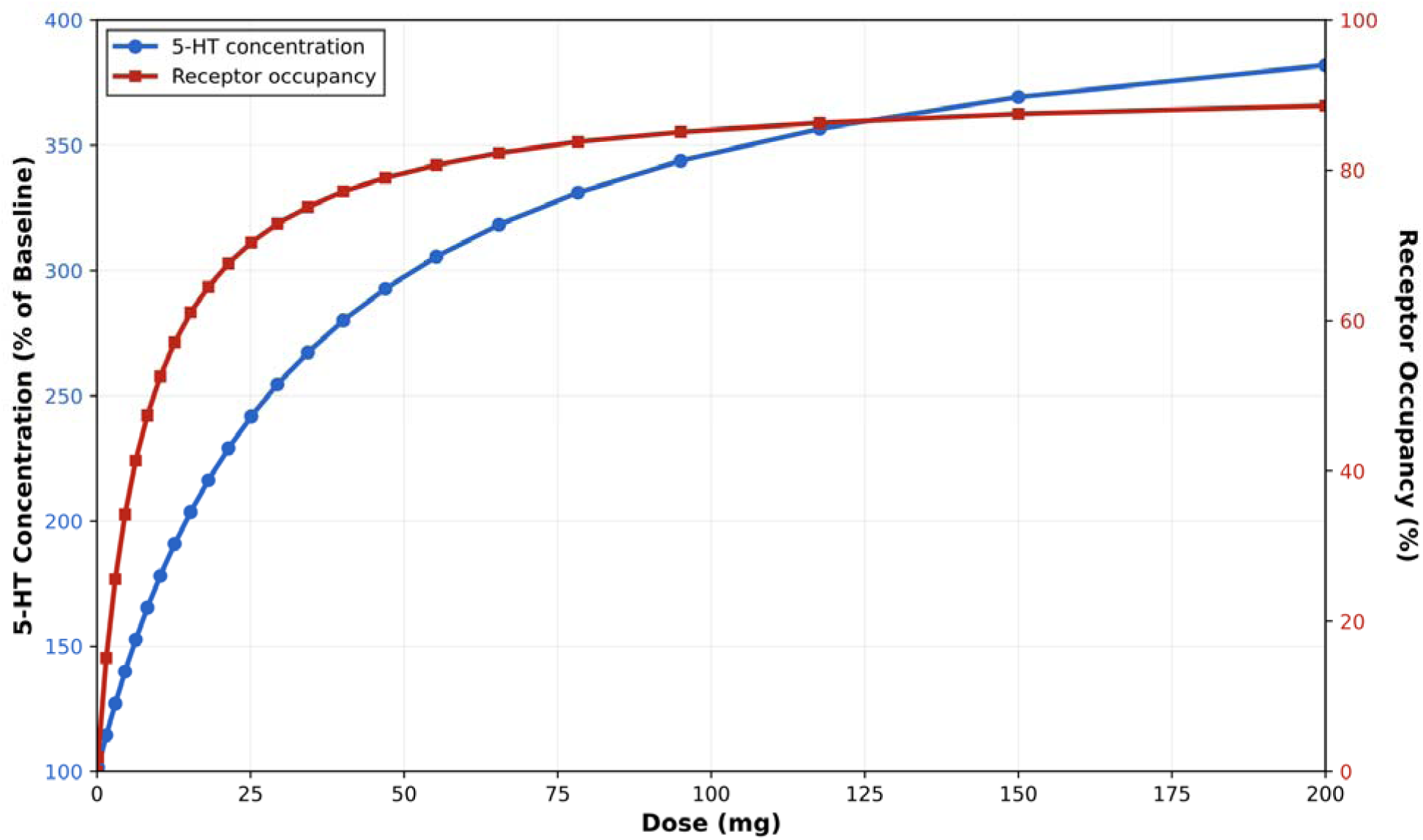
Dose-effect relationships graphed as both a dose-concentration curve (blue line) and as a dose-occupancy curve (red line) for daily doses of sertraline. E_max_ of 92% and ED_50_ of 7.7 mg for sertraline was employed,(Sørensen et al., 2022) consistent with the Maudsley Deprescribing Guidelines.(Taylor and Horowitz, 2024) Graphed points represent a taper maintaining consistent 3.3% relative reductions in extracellular serotonin concentration per step, shown on both the dose–concentration and dose–occupancy curves. Abbreviations: 5-HT = serotonin; Emax = maximum SERT occupancy; ED50 = dose producing 50% maximal occupancy

SRIs are known to exhibit regional differences in SERT occupancy. (Baldinger et al., 2014; Kim et al., 2017) Within our model, these regional differences may carry greater clinical significance, as they translate to greater differences in C_5-HT_ between brain regions. Because distinct brain regions are known to be implicated in various psychiatric disorders (e.g. hyperactive amygdala in anxiety disorders (Etkin and Wager, 2007); hippocampal dysfunction in depressive disorders (Gray et al., 2020)), regional variation in serotonergic effects may represent one mechanism contributing to differential dose-response relationships across disorders. This possibility warrants further investigation.

Applying dose-occupancy relationships for chronic sertraline treatment established in radioligand imaging studies, we can generate taper regimens that maintain stable reductions in C_5-HT_ to mitigate withdrawal risk (Supplementary Table 1). Stable reductions in C_5-HT_ can be either constant relative (percentage of prior dose carried forward) stepwise reductions in C_5-HT_, or constant numerical (absolute) stepwise reductions in C_5-HT_. Mass-action kinetics dictate that, for receptor systems governed by reversible ligand binding, equal absolute changes in ligand concentration can produce larger proportional changes in receptor occupancy at lower concentrations. (Goutelle et al., 2008) Also, several 5-HT receptor subtypes including the 5-HT_1A_ autoreceptor (Yocca et al., 1992) exhibit receptor reserve, meaning maximal biological responses can be achieved at less than full receptor occupancy due to post-receptor signal amplification and excess receptor density, both of which can render the system relatively more sensitive to changes at low occupancy levels. These receptor-level dynamics entail that equal absolute reductions in C_5-HT_ during tapering may produce escalating biological effects as concentrations decrease, suggesting that consistent relative (rather than absolute) reductions in C_5-HT_ are most likely to mitigate risk of withdrawal symptoms.

This model may be used to design taper plans based on any well-tolerated initial dose reduction, with subsequent dose decreases calibrated to maintain a consistent relative change in C_5-HT_. For instance, a patient initiating a taper of sertraline 200 mg daily who tolerates the initial dose reduction to 150 mg (a 3.3% relative reduction in C_5-HT_) can maintain a constant 3.3% C_5-HT_ reduction rate throughout their taper (Supplementary Table 1a), with stepwise reductions at 2-4 week intervals, as tolerated. Figure 4 graphs this taper both in terms of C_5-HT_ and RO. For patients who have not yet started to taper their SRI, an initial reduction of 3-5% in C_5-HT_ may be advisable, with this relative change in C_5-HT_ maintained throughout the taper if the initial step is well-tolerated (Supplementary Table 1b).

### Comparison with existing frameworks

The Maudsley Deprescribing Guidelines have influenced clinical guidance in the UK(Schweitzer and Maguire, 2001) and Australia,(Wilson Foundation, 2024) both of which advocate hyperbolic tapering of antidepressants. In the Maudsley guidelines, taper schedules for SRI antidepressants follow hyperbolic principles in order to “reduce the drug in such a way that produces an even amount of reduction in effect on target receptors.”(Taylor and Horowitz, 2024) However, Maudsley’s recommended taper steps within the therapeutic dosage range do not consistently adhere to fixed, stepwise reductions in RO—likely because doing so would require numerically implausible dose reductions. For example, under Maudsley’s “slower” sertraline taper (described as ∼2.6% stepwise reductions in RO), a 200 mg/day dose is reduced to 150 mg/day (∼1% RO reduction) rather than the predicted ∼108 mg/day (∼2. B 6% RO reduction). For a “faster taper” (10% reductions in RO), a 200 mg/day dose is reduced to 100 mg/day (∼4% RO reduction) rather than to 45 mg/day (∼10% RO reduction).(Taylor and Horowitz, 2024)

For a theoretical sertraline taper (Figure 5), Maudsley’s departures from fixed occupancy reductions in the initial taper steps (1-9) bring the resulting C_5-HT_ reductions closer to the constant relative C_5-HT_ taper that our model predicts, suggesting an implicit attempt to approximate a more consistent biological effect. However, because the Maudsley framework remains grounded in occupancy rather than a variable more proximal to downstream biological activity, it approaches this target inconsistently, resulting in residual variability in C_5-HT_. A fixed-occupancy taper is less erratic, but demonstrates relatively high reductions in C_5-HT_ at initial taper steps and extremely small reductions at later phases (Figure 5). Both of these approaches may underestimate biologically relevant changes occurring at higher doses, and may overestimate them at very low doses. However, our model still predicts the need for cautious tapering to very low doses to maintain a consistent biological effect across taper steps, consistent with current hyperbolic tapering guidance.

**Figure 5:**
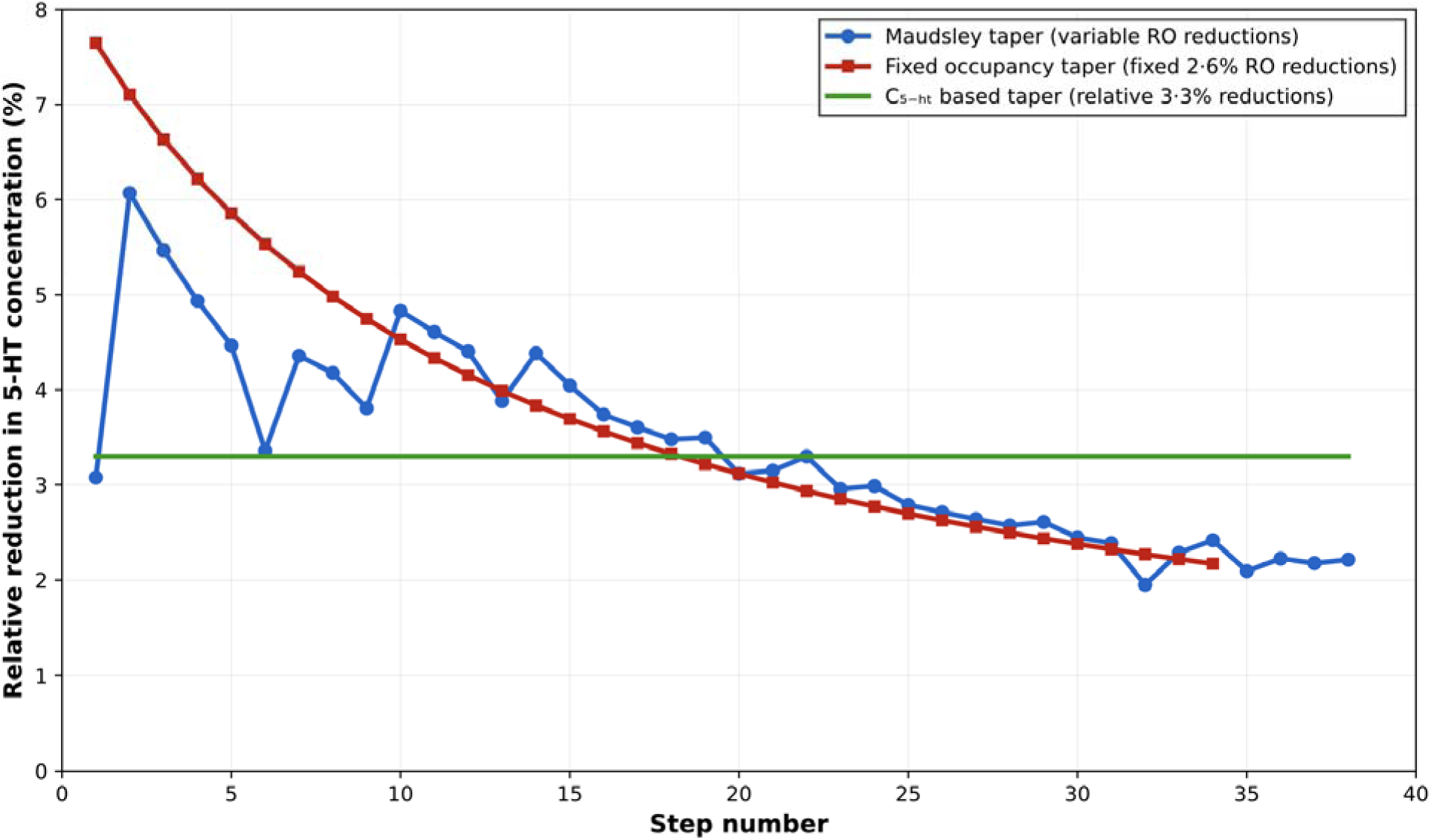
Comparison of the “slow” taper from the MDG (blue line) vs. a taper maintaining consistent 2.6% SERT occupancy (RO) reductions per step (red line) vs. a taper maintaining consistent 3.3% relative reductions in extracellular serotonin concentration (C_5-HT_) per step (green line), for sertraline starting at 200mg/day. Last step excluded from each taper for illustrative purposes. Abbreviations: MDG= Maudsley Deprescribing Guidelines; SERT=serotonin transporter

### Generalization to other transporter systems

The hyperbolic relationship between transporter occupancy and extracellular serotonin concentrations under chronic treatment is ultimately dictated by the same law of mass-action that underlies the hyperbolic SRI dose-SERT occupancy curve. As transporter occupancy increases, free ligands (SRI molecules or C_5-HT_) encounter a progressively smaller pool of available transporters. For example, increasing occupancy from 0% to 10% only reduces the available transporter pool by 10%, whereas increasing occupancy from 80% to 90% reduces it by 50%. Equivalent increases in occupancy therefore correspond to progressively larger reductions in available transporter capacity as occupancy increases, yielding accelerating serotonin concentrations as occupancy increases.

Because this principle is not specific to the serotonergic system, we hypothesize that an analogous relationship may apply to other transporter systems whose occupancy is not directly coupled to biological effect, such as presynaptic norepinephrine and dopamine transporters. If validated, this framework may offer a possible mechanistic explanation for dose-dependent risks of psychostimulants (e.g. cardiac risks or euphoria) and norepinephrine reuptake inhibitors. In contrast, receptor systems that are more directly coupled to biological effect, such as postsynaptic D_2_ receptors or somatodendritic and postsynaptic 5-HT_1A_ receptors, may more closely approximate a linear occupancy-effect relationship and be more appropriately tapered using current hyperbolic approaches based on available dose-occupancy data.

### Implications for serotonin toxicity

Our two-pathway model also provides insight into other clinically relevant phenomena, such as serotonin toxicity. Severe, life-threatening serotonin syndrome occurs almost exclusively when monoamine oxidase inhibitors (MAOis) are combined with potent serotonin reuptake inhibitors.(Gillman, 2005) Because MAO_A_ constitutes the dominant non-SERT pathway for serotonin clearance,(Fitzgerald et al., 1990) near-complete MAO inhibition effectively drives the non-SERT clearance parameter (K_2_) toward zero (Eq. 1). Under conditions of significant MAO inhibition, concurrent SERT blockade can produce dramatic increases in C_5-_HT, precipitating significant serotonin syndrome when levels exceed approximately 10-fold above baseline.(Tao et al., 2014) In contrast, isolated SRI overdoses are predicted by our model to yield C_5-HT_ generally within the 5-9x of baseline range observed in SERT knockout models (Figure 3), and hence, are associated with a low risk of significant serotonin toxicity.

### Comparison to Best *et al*.’s model

Best *et al*.’s differential model was designed to capture detailed temporal and kinetic structure.(Best et al., 2010) Our algebraic formulation, which collapses many of these biological processes into a few effective parameters, does not aim to replicate the mechanistic precision of their framework, but rather to provide a computationally simple mapping from RO to C_5-HT_. This makes it well-suited for applications such as taper modeling where the primary goal is to estimate relative changes in C_5-HT_ rather than simulate full serotonergic dynamics.

Their model was parameterised to reflect the SRI-naïve state, assuming fully intact 5-HT₁A autoreceptors that progressively suppress serotonin release and synthesis as RO increases. Under these assumptions, the absolute increase in C_5-HT_ from 80–90% RO is approximately 92% of that observed from 0–50% RO, substantially less than the ∼141% increase predicted by our model. However, incorporating partial (e.g. 50%) autoreceptor desensitisation into their framework would markedly increase the predicted growth, bringing it into closer agreement with our results.

## 5. Limitations

Several limitations of our model should be considered. Although extracellular serotonin concentrations are more proximal to serotonergic signaling than transporter occupancy alone, downstream changes induced by chronic SRI treatment which are not accounted for in our model include somatodendritic, presynaptic, and postsynaptic serotonin receptor adaptations,(Quentin et al., 2018) SERT downregulation,(Benmansour et al., 2002) and activity-dependent amplification of serotonin release.(Dankoski et al., 2014)

Due to desensitization, 5-HT_1A_ autoreceptors—the principal autoregulatory mechanism limiting serotonin synthesis and release—undergo an estimated 40–50% reduction in function during chronic SRI treatment at therapeutic doses, resulting in increased presynaptic serotonin release.(Pejchal et al., 2002),(Rossi et al., 2008) While our model does not explicitly incorporate autoreceptor feedback, it is anchored to experimental C_5-HT_ values measured after chronic SRI treatment, and therefore implicitly reflects the net effects of autoreceptor desensitization present under these conditions. Because our model conforms to the general form of a hyperbolic clearance function (y=A/(B-x)), it can alternatively be derived purely by fitting a hyperbolic function to two empirical anchors: an elevation to 300-350% of baseline C_5-HT_ at ∼80-85% SERT occupancy after chronic SRI treatment,(Anderson et al., 2005; Fritze et al., 2017) and a theoretical maximum elevation of 600% of baseline, consistent with SERT knockout studies.(Fabre et al., 2000; Gobbi et al., 2001; Homberg et al., 2007) Together, these empirical anchors and the underlying hyperbolic form significantly constrain the shape and magnitude of the RO–C_5-HT_ relationship. As a result, a model that explicitly incorporates autoreceptor feedback would not be expected to diverge substantially from our current model, particularly within the therapeutic dose range. However, the degree of autoreceptor desensitization at lower doses and ROs has not been well characterized, so the precise effects on C_5-HT_ in this range remain an open question.

Our model reproduces both whole-brain and frontal cortical estimates, and the predicted elevation to 6x baseline at theoretical complete transporter blockade (100% RO) represents a reasonable conservative estimate of the average elevation observed across most brain regions assessed in SERT-knockout studies, which generally report elevations to ∼5–9x baseline. (Fabre et al., 2000; Gobbi et al., 2001; Homberg et al., 2007) However, frontal cortical serotonergic elevations may still be underestimated. In this region, the ∼300–350% elevation following chronic SRI treatment was derived from microdialysis studies incorporating a one-day washout period prior to measurement, which would be expected to reduce measured C_5-HT_. (Fritze et al., 2017) The frontal cortex also demonstrates comparatively larger serotonergic elevations than most other brain regions in SERT-knockout studies, approaching ∼10x baseline in some reports (Mathews et al., 2004), suggesting that our model’s predicted 6x baseline at theoretical complete transporter blockade may underestimate frontal cortical effects. Additionally, SERT-knockout models themselves may underestimate the elevation in C_5-HT_ expected under theoretical complete transporter blockade, as developmental compensation in knockout animals tends to attenuate serotonergic elevations (Baganz et al., 2008)

Estimates of C_5-HT_ in our model were based on preclinical animal studies that may not be fully generalizable to humans. Additional uncertainty arises from methodological heterogeneity and limited sample sizes in human radioligand imaging studies characterizing SRI dose–occupancy relationships, as well as the application of striatal occupancy data to approximate whole-brain and frontal cortical dose–occupancy relationships. The degree of C_5-HT_ elevation after chronic SRI treatment also varies significantly by brain region(Fritze et al., 2017). While these factors introduce uncertainty into the precise absolute levels of extracellular serotonin predicted by our model, the qualitative hyperbolic relationship between transporter occupancy and C_5-HT_ under chronic treatment follows from mass-action kinetics and does not depend on the specific parameter values chosen.

Multiple risk factors for SRI withdrawal have previously been identified, including longer treatment duration, short elimination half-life, and prior withdrawal symptoms.(Horowitz et al., 2023) Our model provides a potential mechanistic explanation for higher therapeutic doses as an additional risk factor.(Horowitz et al., 2023) However, substantial uncertainty remains regarding why some individuals require prolonged hyperbolic tapers to ultra-low doses, whereas others tolerate linear tapering and discontinue at the minimum available dose without clinically significant withdrawal. These observations suggest that withdrawal vulnerability is likely shaped by factors beyond serotonergic concentration changes alone. Future research should further identify determinants of withdrawal sensitivity and the need for slower tapers or tapering to ultra-low doses.

Individual differences in SERT expression, monoamine oxidase activity, cytochrome P450 (CYP)-mediated SRI metabolism, 5-HT autoreceptor function, and serotonin synthesis and release may alter the shape of the dose–C_5-HT_ relationship and thereby influence withdrawal risk. Modeling these effects represents an important future research direction that may generate testable predictions regarding individual vulnerability to withdrawal. Quantitative modeling of downstream 5-HT receptor dynamics is beyond the scope of the present analysis but may represent an additional avenue for future investigation. Clinically, controlled studies comparing SRI tapering strategies are warranted. Preclinical microdialysis studies assessing extracellular serotonin concentrations at lower SRI doses are also needed.

## 6. Conclusion

Our two-pathway kinetic model predicts a hyperbolic relationship between SERT occupancy and extracellular serotonin concentrations, challenging the notion that SERT occupancy correlates linearly with biological effect. Its findings provide a potential mechanistic explanation for dose–side-effect and dose–response trends associated with SRI antidepressant treatment that are not readily accounted for by existing occupancy-based models, with implications that may extend to other classes of psychiatric drugs and other important clinical phenomena related to drug toxicity. Importantly, our model provides a novel framework for informing antidepressant tapering strategies, though empirical validation is warranted.

## Data Availability

All data produced in the present work are contained in the manuscript

## Declarations

### Author contributions

Both authors contributed to all aspects of this study: each conceived and designed the study, developed the model, conducted the analyses, interpreted the findings, and drafted the manuscript. Both authors approved the final version of the manuscript.

### Data availability statement

An interactive web-based calculator implementing the model described in this study is available at https://psychfiles.github.io/SRI-taper-calculator-/. The tool is provided for transparency and educational purposes and allows users to explore predicted relative changes in extracellular serotonin across dose reductions, including dose sequences that maintain consistent relative reductions in extracellular serotonin concentration as described in this paper. It has not been validated against clinical outcomes.

## Bibliography

Anderson GM, Barr CS, Lindell S, et al. (2005) Time course of the effects of the serotonin-selective reuptake inhibitor sertraline on central and peripheral serotonin neurochemistry in the rhesus monkey. Psychopharmacology 178(2–3): 339–346. DOI: 10.1007/s00213-004-2011-7.

Baganz NL, Horton RE, Calderon AS, et al. (2008) Organic cation transporter 3: Keeping the brake on extracellular serotonin in serotonin-transporter-deficient mice. Proceedings of the National Academy of Sciences of the United States of America 105(48): 18976–18981. DOI: 10.1073/pnas.0800466105.

Baldinger P, Kranz GS, Haeusler D, et al. (2014) Regional differences in SERT occupancy after acute and prolonged SSRI intake investigated by brain PET. Neuroimage 88: 252–262. DOI: 10.1016/j.neuroimage.2013.10.002.

Benmansour S, Owens WA, Cecchi M, et al. (2002) Serotonin clearance in vivo is altered to a greater extent by antidepressant-induced downregulation of the serotonin transporter than by acute blockade of this transporter. The Journal of Neuroscience 22(15): 6766–6772. DOI: 10.1523/JNEUROSCI.22-15-06766.2002.

Best J, Nijhout HF and Reed M (2010) Serotonin synthesis, release and reuptake in terminals: a mathematical model. Theoretical Biology & Medical Modelling 7: 34. DOI: 10.1186/1742-4682-7-34.

Bloch MH, McGuire J, Landeros-Weisenberger A, et al. (2010) Meta-analysis of the dose-response relationship of SSRI in obsessive-compulsive disorder. Molecular Psychiatry 15(8): 850–855. DOI: 10.1038/mp.2009.50.

Braun C, Adams A, Rink L, et al. (2020) In search of a dose–response relationship in SSRIs: a systematic review, meta-analysis, and network meta-analysis. Acta Psychiatrica Scandinavica 142(6): 430–442. DOI: 10.1111/acps.13235.

Dankoski EC, Agster KL, Fox ME, et al. (2014) Facilitation of serotonin signaling by SSRIs is attenuated by social isolation. Neuropsychopharmacology 39(13): 2928–2937. DOI: 10.1038/npp.2014.162.

Etkin A and Wager TD (2007) Functional neuroimaging of anxiety: a meta-analysis of emotional processing in PTSD, social anxiety disorder, and specific phobia. The American Journal of Psychiatry 164(10): 1476–1488. DOI: 10.1176/appi.ajp.2007.07030504.

Fabre V, Beaufour C, Evrard A, et al. (2000) Altered expression and functions of serotonin 5-HT_1A_ and 5-HT_1B_ receptors in knock-out mice lacking the 5-HT transporter. European Journal of Neuroscience 12(7): 2299–2310. DOI: 10.1046/j.1460-9568.2000.00126.x.

Ferguson JM (2001) SSRI antidepressant medications: adverse effects and tolerability. Primary care companion to the Journal of clinical psychiatry 3(1): 22–27. DOI: 10.4088/pcc.v03n0105.

Fitzgerald LW, Kaplinsky L and Kimelberg HK (1990) Serotonin metabolism by monoamine oxidase in rat primary astrocyte cultures. Journal of Neurochemistry 55(6): 2008–2014. DOI: 10.1111/j.1471-4159.1990.tb05789.x.

Fritze S, Spanagel R and Noori HR (2017) Adaptive dynamics of the 5-HT systems following chronic administration of selective serotonin reuptake inhibitors: a meta-analysis. Journal of Neurochemistry 142(5): 747–755. DOI: 10.1111/jnc.14114.

Furukawa TA, Cipriani A, Cowen PJ, et al. (2019) Optimal dose of selective serotonin reuptake inhibitors, venlafaxine, and mirtazapine in major depression: a systematic review and dose-response meta-analysis. The Lancet. Psychiatry 6(7): 601–609. DOI: 10.1016/S2215-0366(19)30217-2.

Gillman PK (2005) Monoamine oxidase inhibitors, opioid analgesics and serotonin toxicity. British Journal of Anaesthesia 95(4): 434–441. DOI: 10.1093/bja/aei210.

Gobbi G, Murphy DL, Lesch K, et al. (2001) Modifications of the serotonergic system in mice lacking serotonin transporters: an in vivo electrophysiological study. The Journal of Pharmacology and Experimental Therapeutics 296(3): 987–995.

Goutelle S, Maurin M, Rougier F, et al. (2008) The Hill equation: a review of its capabilities in pharmacological modelling. Fundamental & clinical pharmacology 22(6): 633–648. DOI: 10.1111/j.1472-8206.2008.00633.x.

Gray JP, Müller VI, Eickhoff SB, et al. (2020) Multimodal Abnormalities of Brain Structure and Function in Major Depressive Disorder: A Meta-Analysis of Neuroimaging Studies. The American Journal of Psychiatry 177(5): 422–434. DOI: 10.1176/appi.ajp.2019.19050560.

Hagan CE, Neumaier JF and Schenk JO (2010) Rotating disk electrode voltammetric measurements of serotonin transporter kinetics in synaptosomes. Journal of Neuroscience Methods 193(1): 29–38. DOI: 10.1016/j.jneumeth.2010.08.009.

Homberg JR, Olivier JDA, Smits BMG, et al. (2007) Characterization of the serotonin transporter knockout rat: a selective change in the functioning of the serotonergic system. Neuroscience 146(4): 1662–1676. DOI: 10.1016/j.neuroscience.2007.03.030.

Horowitz MA and Taylor D (2019) Tapering of SSRI treatment to mitigate withdrawal symptoms. The Lancet. Psychiatry 6(6): 538–546. DOI: 10.1016/S2215-0366(19)30032-X.

Horowitz MA, Framer A, Hengartner MP, et al. (2023) Estimating Risk of Antidepressant Withdrawal from a Review of Published Data. CNS Drugs 37(2): 143–157. DOI: 10.1007/s40263-022-00960-y.

Horowitz M and Wilcock M (2025) Hyperbolic tapering of antidepressants: where are we now? Drug and Therapeutics Bulletin 64(1): dtb-2024-000065. DOI: 10.1136/dtb.2024.000065.

Jakubovski E, Varigonda AL, Freemantle N, et al. (2016) Systematic review and meta-analysis: dose-response relationship of selective serotonin reuptake inhibitors in major depressive disorder. The American Journal of Psychiatry 173(2): 174–183. DOI: 10.1176/appi.ajp.2015.15030331.

Jakubovski E, Johnson JA, Nasir M, et al. (2019) Systematic review and meta-analysis: Dose-response curve of SSRIs and SNRIs in anxiety disorders. Depression and Anxiety 36(3): 198–212. DOI: 10.1002/da.22854.

Kendrick T (2021) Strategies to reduce use of antidepressants. British Journal of Clinical Pharmacology 87(1): 23–33. DOI: 10.1111/bcp.14475.

Kim E, Howes OD, Kim B-H, et al. (2017) Regional Differences in Serotonin Transporter Occupancy by Escitalopram: An [11C]DASB PK-PD Study. Clinical Pharmacokinetics 56(4): 371–381. DOI: 10.1007/s40262-016-0444-x.

Mathews TA, Fedele DE, Coppelli FM, et al. (2004) Gene dose-dependent alterations in extraneuronal serotonin but not dopamine in mice with reduced serotonin transporter expression. Journal of Neuroscience Methods 140(1–2): 169–181. DOI: 10.1016/j.jneumeth.2004.05.017.

Meyer JH, Wilson AA, Sagrati S, et al. (2004) Serotonin transporter occupancy of five selective serotonin reuptake inhibitors at different doses: an [11C]DASB positron emission tomography study. The American Journal of Psychiatry 161(5): 826–835. DOI: 10.1176/appi.ajp.161.5.826.

Pejchal T, Foley MA, Kosofsky BE, et al. (2002) Chronic fluoxetine treatment selectively uncouples raphe 5-HT(1A) receptors as measured by [(35)S]-GTP gamma S autoradiography. British Journal of Pharmacology 135(5): 1115–1122. DOI: 10.1038/sj.bjp.0704555.

Quentin E, Belmer A and Maroteaux L (2018) Somatodendritic regulation of raphe serotonin neurons: a key to antidepressant action. Frontiers in Neuroscience 12: 982. DOI: 10.3389/fnins.2018.00982.

Rossi DV, Burke TF, McCasland M, et al. (2008) Serotonin-1A receptor function in the dorsal raphe nucleus following chronic administration of the selective serotonin reuptake inhibitor sertraline. Journal of Neurochemistry 105(4): 1091–1099. DOI: 10.1111/j.1471-4159.2007.05201.x.

Rowland M and Tozer TN (2019) Clinical Pharmacokinetics and Pharmacodynamics: Concepts and Applications. 5th ed. Philadelphia, PA: Wolters Kluwer.

Schweitzer I and Maguire K (2001) Stopping antidepressants. Australian prescriber 24(1): 13–15. DOI: 10.18773/austprescr.2001.008.

Shapiro BB (2018) Subtherapeutic doses of SSRI antidepressants demonstrate considerable serotonin transporter occupancy: implications for tapering SSRIs. Psychopharmacology 235(9): 2779–2781. DOI: 10.1007/s00213-018-4995-4.

Sørensen A, Ruhé HG and Munkholm K (2022) The relationship between dose and serotonin transporter occupancy of antidepressants: a systematic review. Molecular Psychiatry 27(1): 192–201. DOI: 10.1038/s41380-021-01285-w.

Tao R, Ma Z and Auerbach SB (2000) Differential effect of local infusion of serotonin reuptake inhibitors in the raphe versus forebrain and the role of depolarization-induced release in increased extracellular serotonin. The Journal of Pharmacology and Experimental Therapeutics 294(2): 571–579.

Tao R, Rudacille M, Zhang G, et al. (2014) Changes in intensity of serotonin syndrome caused by adverse interaction between monoamine oxidase inhibitors and serotonin reuptake blockers. Neuropsychopharmacology 39(8): 1996–2007. DOI: 10.1038/npp.2014.49.

Taylor D and Horowitz M (2024) The Maudsley Deprescribing Guidelines: Antidepressants, Benzodiazepines, Gabapentinoids and Z-Drugs. Hoboken, NJ: Wiley. DOI: 10.1002/9781394291052.

US Food and Drug Administration (2024) Lexapro (escitalopram) tablets prescribing information. Available at: https://www.accessdata.fda.gov/drugsatfda_docs/label/2024/021323s058lbl.pdf (accessed 23 January 2026).

Verleysdonk S, Hamprecht B, Rapp M, et al. (2004) Uptake and metabolism of serotonin by ependymal primary cultures. Neurochemical Research 29(9): 1739–1747. DOI: 10.1023/b:nere.0000035810.08543.97.

Ward W, Haslam A and Prasad V (2025) Antidepressant trial duration versus duration of real-world use: a systematic analysis. The American Journal of Medicine 138(10): 1400–1407.e10. DOI: 10.1016/j.amjmed.2025.04.037.

Wilson Foundation (2024) Royal Australian College of General Practitioners endorses landmark safe deprescribing guide. Available at: https://www.wilsonfoundation.org.au/news/royal-australian-college-of-general-practitioners-endorses-landmark-safe-deprescribing-guide (accessed 20 January 2026).

Yocca FD, Iben L and Meller E (1992) Lack of apparent receptor reserve at postsynaptic 5-hydroxytryptamine1A receptors negatively coupled to adenylyl cyclase activity in rat hippocampal membranes. Molecular Pharmacology 41(6): 1066–1072.

