## Supplementary Table for "The relationship between serotonin transporter occupancy and extracellular serotonin concentration is hyperbolic, not linear: implications for safely tapering antidepressants"

a)

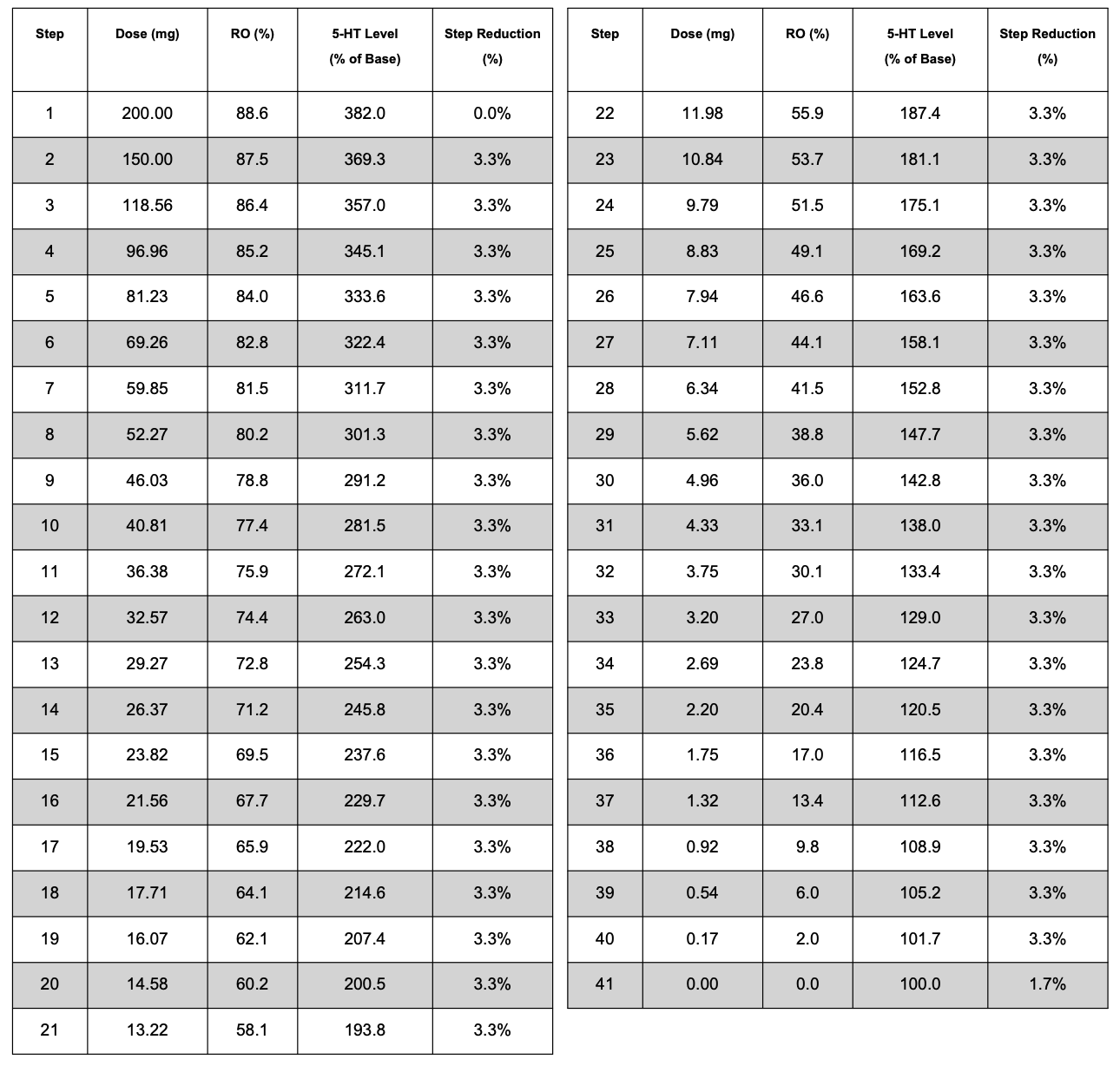

(b)

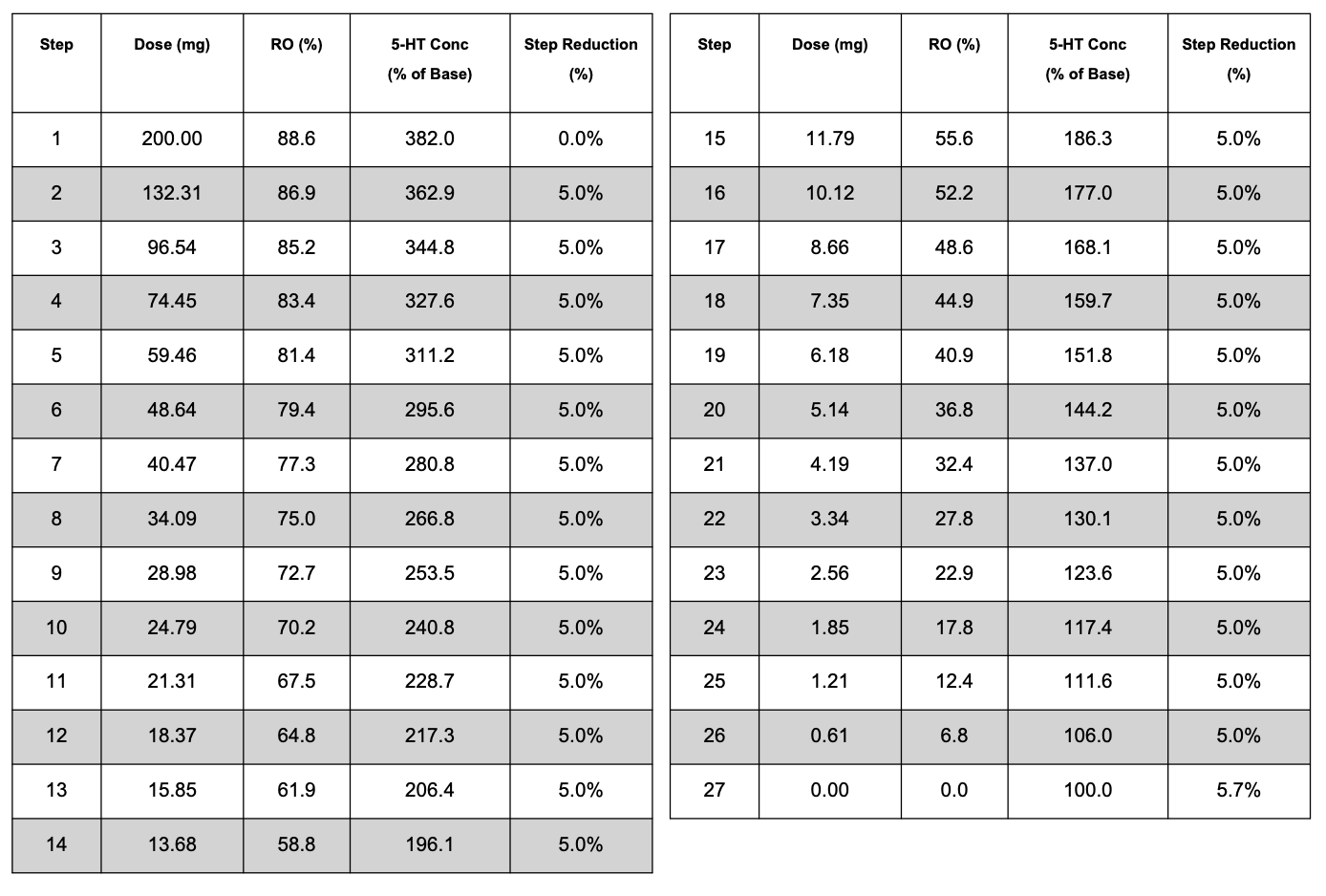

*Supplementary Table 1. Sample taper plans for sertraline incorporating fixed relative reductions in extracellular 5-HT concentrations, with 3.3% stepwise reductions in extracellular 5-HT concentrations (Panel A) and 5% stepwise reductions in extracellular 5-HT concentrations (Panel B).*
